# USING SPUTUM AND TONGUE SWAB SPECIMENS FOR IN-HOME POINT-OF-CARE TARGETED UNIVERSAL TESTING FOR TB OF HOUSEHOLD CONTACTS: AN ACCEPTABILITY AND FEASIBILITY ANALYSIS

**DOI:** 10.1101/2024.11.01.24316570

**Authors:** Charl Bezuidenhout, Lawrence Long, Brooke Nichols, Gesine Meyer-Rath, Matthew P Fox, Sharon Olifant, Grant Theron, Kuhle Fiphaza, Morten Ruhwald, Adam Penn-Nicholson, Bernard Fourie, Andrew Medina-Marino

## Abstract

**Introduction:** Effective strategies are needed to facilitate early detection and diagnosis of tuberculosis (TB). The over-reliance on passive case detection, symptom screening, and collection of sputum, results in delayed or undiagnosed TB, which directly contributes to on-going TB transmission. We assessed the acceptability and feasibility of in-home, Targeted Universal TB Testing (TUTT) of household contacts using GeneXpert MTB/RIF Ultra at point-of-care (POC) during household contact investigations (HCIs) and compared the feasibility of using sputum vs. tongue swab specimens.

**Methods:** Household contacts (HHCs) receiving in-home POC TUTT as part of the TB Home Study were asked to complete a post-test acceptability survey. The survey explored HHC’s level of comfort, confidence in the test results, and the perceived appropriateness of in-home POC TUTT. We used the Metrics to Assess the Feasibility of Rapid Point-of-Care Technologies framework to assess the feasibility of using sputum and tongue swab specimens for in-home POC TUTT. Descriptive statistics were used to report participant responses and feasibility metrics.

**Results:** Of 313 eligible HHCs, 267/313 (85.3%) consented to in-home POC TUTT. Of those, 267/267 (100%) provided a tongue swab and 46/267 (17.2%) could expectorate sputum. All specimens were successfully prepared for immediate, in-home testing with Xpert Ultra on GeneXpert Edge. Of 164 tongue swab tests conducted, 160/164 (97.6%) generated a valid test result compared to 44/46 (95.7%) sputum-based tests. An immediate test result was available for 262/267 (98.1%) individuals based on in-home swab testing, and 44/46 (95.7%) based on in-home sputum testing. The mean in-home POC TUTT acceptability score (5=highly acceptable) was 4.5/5 (SD= 0.2).

**Conclusion:** In-home, POC TUTT using either sputum or tongue swab specimens was highly acceptable and feasible. Tongue swab specimens greatly increase the proportion of HHCs tested compared to sputum. In-home POC TUTT using a combination of sputum and tongue swabs can mitigate shortcomings to case detection.

**KEY MESSAGE:** 

**What is already known on this topic:**

- TB transmission among household contacts of people with TB is a public health concern.
- The delivery of community-based diagnostic testing for TB is challenging and the reliance on sputum continue to hamper universal testing and result in diagnostic delay.

**What this study adds:**

- This is the first study to assess the acceptability of universal in-home point-of-care TB testing of household contacts during household contact investigations.
- This study assesses the feasibility of different specimen types for immediate in-home point-of-care TB testing including tongue swabs and sputum.

**How this study might affect research, practice or policy:**

- Household contacts perceived in-home targeted universal TB testing to be highly acceptable, prompting the need for further investigation into the cost-effectiveness of such strategies to improve early case detection.
- The use of tongue swabs as an additional or alternative sample type to sputum could increase testing and improve early case detection.

## INTRODUCTION

Tuberculosis (TB) remains one of the world’s leading infectious disease killers. Despite being preventable, treatable, and curable it caused an estimated 1.3 million deaths in 2022.^1^ Persistent gaps in the cascade of care include TB diagnosis, notification, and linkage to treatment, all of which remain major contributors to TB burden, transmission, and mortality.^2^ Early screening and diagnosis with World Health Organization (WHO)-recommended rapid molecular diagnostic tests like GeneXpert MTB/RIF Ultra (Cepheid, Sunnyvale, CA, USA) (Xpert Ultra) remains a key priority to meet the WHO End TB goal to achieve an 80% reduction in the annual TB deaths by 2030.^1^ Rapid diagnosis followed by immediate treatment initiation is crucial to eradicating TB.^3^ However, despite the introduction of rapid molecular diagnostic tests like Xpert Ultra, diagnosis remains the weakest link in the cascade of care.^4^

In 2021, South Africa’s TB incidence was estimated to be 513 per 100,000 population, equating to ~304,000 people living with TB, of which ~120,000 were not tested, diagnosed, or initiated on treatment.^5,6^ Persistent gaps in testing are attributable to a variety of factors including reliance on symptoms, symptom screening, passive case detection, limited access to testing, and fragmented delivery of testing services that requires clients to return for multiple clinic visits.^7–10^ Closing these gaps necessitates strategies that are patient-centered, deliver testing at or near the point-of-care (POC), are conducted in a single patient consultation, and are consistently provided to high risk groups.^4,11^

Active case finding strategies, including household contact investigation (HCI) of people diagnosed with TB, have shown to be cost-effective compared to passive case detection and are increasingly recognized as a cornerstone of TB programs aimed at improving early case detection.^12–14^ However, low uptake of clinic referrals, long clinic waiting times, and continued reliance on a hub-and-spoke model of sputum transportation and centralized testing (resulting in long test result turnaround) have all been cited as challenges hampering effective implementation of such strategies.^15^ To address some of these limitations, we previously explored the acceptability and feasibility of using the GeneXpert Edge (GX-Edge) for in-home, Xpert Ultra POC testing of symptomatic household contacts (HHCs) as part of HCI. This adapted HCI strategy was acceptable and feasible, improved the proportion of symptomatic HHCs tested, and reduced test notification turnaround time, compared to those referred for clinic-based testing.^8,16^ Despite the relative success, reliance on symptom-based screening and sputum production limited case detection.^17^

Non-specific clinical symptoms, the paucibacillary nature of sputum, and the challenge of collecting induced or expectorated sputum have all been reported as barriers to effective diagnostic testing.^18,19^ Furthermore, the high cost of Xpert testing as part of community-based active case finding strategies severely limits its scalability in resource constrained settings with a high TB burden.^20^ However, significant strides have been made to overcome both these barriers. Specifically, using tongue swab specimens as an alternative, less invasive sample, when sputum is not available has received increased attention, especially as sensitivity approaches that of sputum-based molecular tests.^21^ Moreover, pooled testing of multiple samples in a single cartridge on Xpert Ultra has shown to save up to 48% of assay cost.^22^ Combined, the collection of tongue swabs to increase sample yield and the pooling of samples to decrease cost may increase the likelihood of scalability of in-home POC testing during HCI.^14^ Given the introduction of targeted universal testing for TB (TUTT) of all HHCs, irrespective of symptoms, there is urgent need for rapid, affordable, and accurate TB screening (and testing) strategies that could be applied universally.^11^

Exploration of the acceptability and feasibility of in-home, POC TUTT using tongue swab specimens is warranted. In order to make prudent decisions about adopting new technologies, decision-makers need well-executed studies assessing their acceptability and feasibility.^23^ Just because a rapid test is easy to perform does not mean it’s easy to implement, especially at POC.^24^ A variety of contextual factors and patient preferences could ultimately determine the success of new healthcare interventions. It’s imperative to explore and understand dynamics at the level of intended use.^25^ To this end, we sought to: 1) assess the acceptability of in-home, POC TUTT of HHCs using the GX-Edge; and 2) to compare the test feasibility of using sputum vs. tongue swab specimens.

## METHODS

### Study Design

This acceptability and feasibility assessment was nested within the larger TB Home Study. Data for this assessment were collected between March and September 2024. The TB Home Study sought to evaluate the predictive value of individual and pooled tongue swab specimens vs. sputum as a household-level triage test for TB during HCIs.^26^ In brief, individuals with microbiologically confirmed TB were asked for permission to visit their homes and conduct a HCI. All consenting HHCs were asked to provide both a sputum and tongue swab for immediate, in-home TB testing using the GX-Edge platform with Xpert Ultra. All HHCs were asked to complete an acceptability survey following the completion of in-home testing. The Theoretical Framework of Acceptability (TFA) guided the development of survey items designed to assess HHCs’ experiences with in-home POC TB testing.^27^ The Metrics to Assess the Feasibility of Rapid Point-of-Care Technologies framework was used to assess implementation outcomes associated with the feasibility of using sputum vs. tongue swab samples during in-home POC TUTT.^28^

### Study setting

The TB Home Study was conducted in the Buffalo City Metro (BCM) Health District, Eastern Cape Province, South Africa. BCM has a population size of approximately 893,000 of which 86.7% are Black. An estimated 45.3% of households are headed by women, and 24.9% of households reside in informal dwellings. An estimated 58.2% of people live in poverty and 31.1% remain unemployed.^29^ In 2019, BCM had an estimated TB incidence of 876 per 100,000 population.^30^ In 2018, the last year in which data are available, BCM had a drug-susceptible TB (DS-TB) treatment success rate of 71.2% (the lowest in South Africa), a loss-to-care rate of 17.6% (second highest in South Africa), and an estimated 40% of TB cases missed by the health system.^31,32^ TB is the leading cause of death (18.4%) amongst the 25-64 age group in BCM.^29^

### Household Contact Recruitment

Details regarding the HCI methods have been previously described.^16,33^ Briefly, household and HHC information was obtained from people with microbiologically confirmed TB accessing services at collaborating public healthcare clinics. Contact investigation teams consisting of 2-3 trained lay community healthcare workers made up to three household visits to reach all HHCs listed. During each household visit, the contact investigation team would conduct a HHC verification check, screen for study eligibility, and introduce the study to all those present. HHCs were deemed eligible if they were: 1) age ≥18 years; 2) not currently on TB treatment; and 3) willing to provide informed consent. Once recruited into the study, participating HHCs were asked to respond to a series of survey questions as well as provide specimens for immediate in-home testing.

### Specimen Collection and Testing

Details regarding the sample collection and testing have been previously reported.^16,26,34–36^ Briefly, prior to sputum collection, Copan FLOQSwabs were used to collect tongue swab specimens from all study participants present at the time of the HCI. Tongue swabs were pooled from up to three individuals for immediate in-home testing using a single Xpert Ultra cartridge. If a household had more than three HHCs, the additional swabs were pooled and tested in a separate reaction. Pool sizes could be three, two, or in some cases a single swab. Sputum samples were collected from all study participants while the tongue swab test was being conducted. Participants unable to expectorate sputum were offered sputum induction using a nebulizer. Those still unable to expectorate sputum were referred to a clinic for further clinical evaluation. Sputum samples were individually prepared and tested immediately in the house using Xpert Ultra with the GX-Edge platform. Testing took ~90 minutes. Participants were referred for TB treatment based on positive sputum results.

### Data Collection and Analysis

While Xpert testing was being conducted, a contact investigation team member collected basic socio-demographic and clinical history data from each participant. Descriptive statistics (median [IQR] for continuous variables and n [%] for categorical variables) were used to characterize distributions of study variables in the sociodemographic questionnaire. Data collected pre- and post-testing were analyzed to assess the acceptability of in-home, POC TUTT. Pre-test acceptability was assessed as the proportion of HHCs consenting to participate out of the eligible population to whom study participation was offered. Post-test acceptability was assessed using survey data collected following the conclusion of a household investigation. The development of the post-test acceptability survey was informed by the TFA framework, and adapted specifically for this study.^27,37^ This survey evaluated a respondent’s level of acceptability of a healthcare intervention across eight different constructs using a 5-point Likert scale (detailed in Table 2).

**Table 1:**
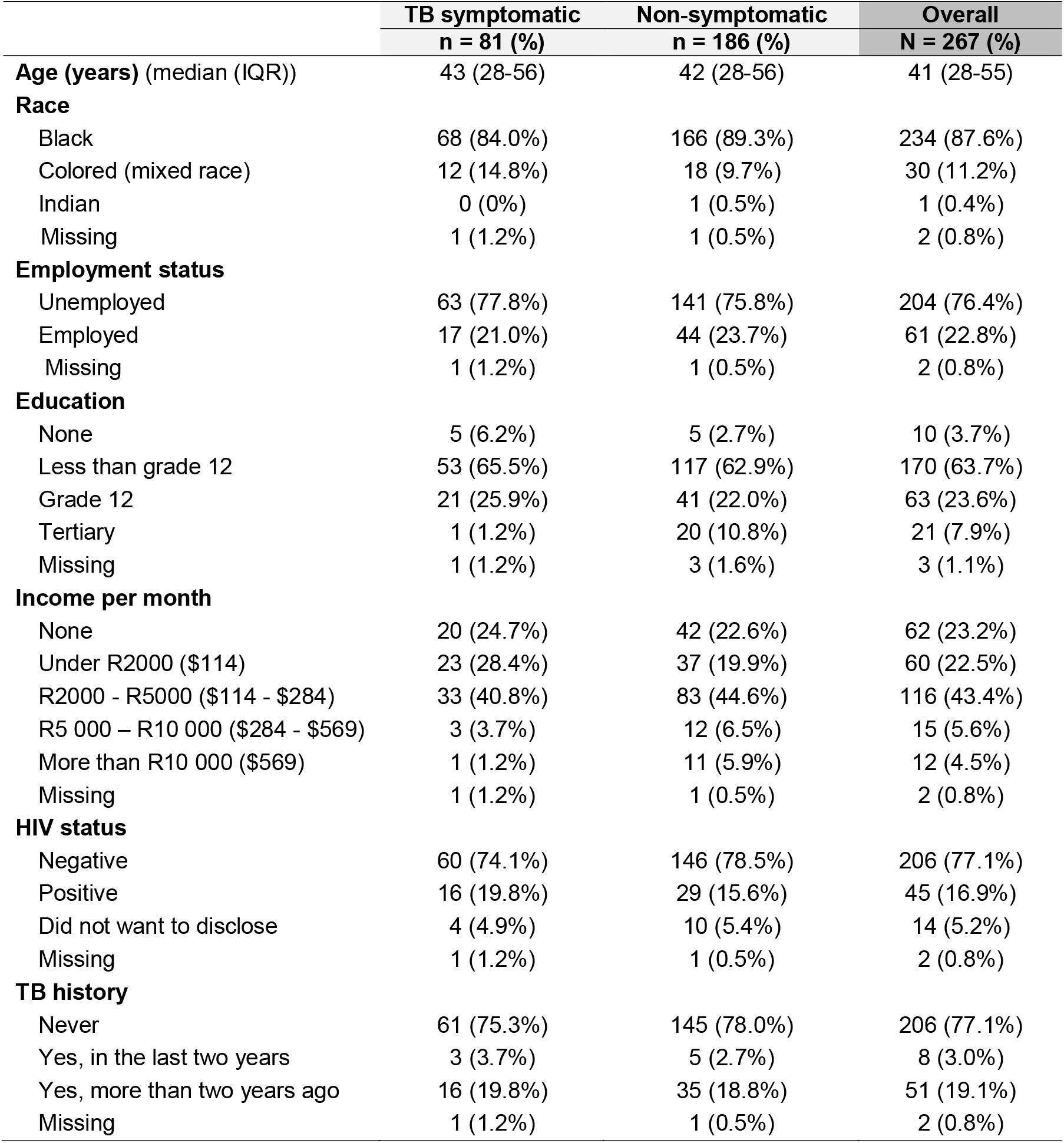
Characteristics of Household Contacts who Received In-Home, POC TUTT and Completed the Post-Test Acceptability Survey.

|  | **TB symptomatic** | **Non-symptomatic** | **Overall** |
| --- | --- | --- | --- |
|  | **n = 81 (%)** | **n = 186 (%)** | **N = 267 (%)** |
| **Age (years)** (median (IQR)) | 43 (28-56) | 42 (28-56) | 41 (28-55) |
| **Race** |  |  |  |
| Black | 68 (84.0%) | 166 (89.3%) | 234 (87.6%) |
| Colored (mixed race) | 12 (14.8%) | 18 (9.7%) | 30 (11.2%) |
| Indian | 0 (0%) | 1 (0.5%) | 1 (0.4%) |
| Missing | 1 (1.2%) | 1 (0.5%) | 2 (0.8%) |
| **Employment status** |  |  |  |
| Unemployed | 63 (77.8%) | 141 (75.8%) | 204 (76.4%) |
| Employed | 17 (21.0%) | 44 (23.7%) | 61 (22.8%) |
| Missing | 1 (1.2%) | 1 (0.5%) | 2 (0.8%) |
| **Education** |  |  |  |
| None | 5 (6.2%) | 5 (2.7%) | 10 (3.7%) |
| Less than grade 12 | 53 (65.5%) | 117 (62.9%) | 170 (63.7%) |
| Grade 12 | 21 (25.9%) | 41 (22.0%) | 63 (23.6%) |
| Tertiary | 1 (1.2%) | 20 (10.8%) | 21 (7.9%) |
| Missing | 1 (1.2%) | 3 (1.6%) | 3 (1.1%) |
| **Income per month** |  |  |  |
| None | 20 (24.7%) | 42 (22.6%) | 62 (23.2%) |
| Under R2000 (\$114) | 23 (28.4%) | 37 (19.9%) | 60 (22.5%) |
| R2000 - R5000 (\$114 - \$284) | 33 (40.8%) | 83 (44.6%) | 116 (43.4%) |
| R5 000 – R10 000 (\$284 - \$569) | 3 (3.7%) | 12 (6.5%) | 15 (5.6%) |
| More than R10 000 (\$569) | 1 (1.2%) | 11 (5.9%) | 12 (4.5%) |
| Missing | 1 (1.2%) | 1 (0.5%) | 2 (0.8%) |
| **HIV status** |  |  |  |
| Negative | 60 (74.1%) | 146 (78.5%) | 206 (77.1%) |
| Positive | 16 (19.8%) | 29 (15.6%) | 45 (16.9%) |
| Did not want to disclose | 4 (4.9%) | 10 (5.4%) | 14 (5.2%) |
| Missing | 1 (1.2%) | 1 (0.5%) | 2 (0.8%) |
| **TB history** |  |  |  |
| Never | 61 (75.3%) | 145 (78.0%) | 206 (77.1%) |
| Yes, in the last two years | 3 (3.7%) | 5 (2.7%) | 8 (3.0%) |
| Yes, more than two years ago | 16 (19.8%) | 35 (18.8%) | 51 (19.1%) |
| Missing | 1 (1.2%) | 1 (0.5%) | 2 (0.8%) |

**Table 2:**
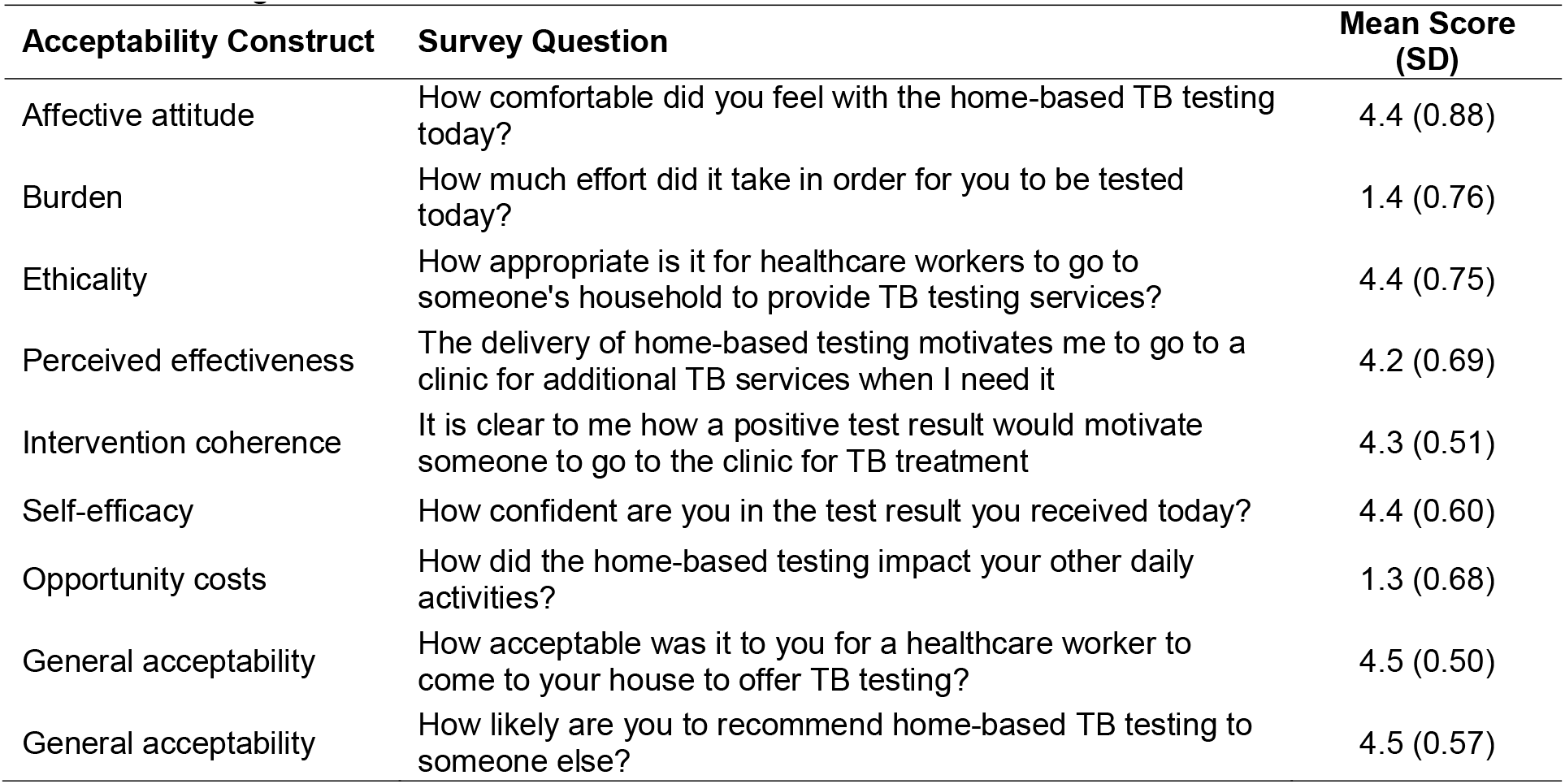
Acceptability of In-home, POC TUTT Among Household Contacts during Household Contact Investigation.

Implementation outcomes associated with sample collection and testing of sputum and tongue swab samples were captured and used to assess the feasibility of different testing methods and included: 1) type of sample collected; 2) success of sample collection; 3) processing and preparation of sample for testing; 4) outcome of test; and 5) referral outcome. The final feasibility assessment was guided by elements of the Metrics to Assess the Feasibility of Rapid Point-of-Care Technologies framework, which was designed to assess the feasibility of rapid POC technologies during proof of concept studies.^38^ This framework was used to select and define four metrics (Table 3) to be included in the current assessment, including: 1) Sample Collection Rate; 2) Test Processing Rate; 3) Test Success Rate; and 4) Patient Notification Rate. The performance of both sputum and tongue swab testing methods against each metric was calculated and reported.

**Table 3:** Performance of sputum and tongue swab specimens during in-home, POC TUTT.

| Metric | Definition | Sputum | Tongue Swab |  |  |  |
| --- | --- | --- | --- | --- | --- | --- |
|  |  | Total | Total | Single Swab | Two Pooled Swabs | Three Pooled Swabs |
| Sample Collection Rate | The proportion of participants able to provide a sample for testing | 46/267<br>(17.2%) | 267/267<br>(100%) | 82/82<br>(100%) | 122/122<br>(100%) | 63/63<br>(100%) |
| Test Processing Rate | The proportion of tests for which all samples were prepared and processed successfully to conduct testing | 46/46<br>(100%) | 267/267<br>(100%) | 82/82<br>(100%) | 122/122<br>(100%) | 63/63<br>(100%) |
| Test Success Rate | The proportion of tests that produced an actionable test result | 44/46<br>(95.7%) | 160/164<br>(97.6%) | 79/82<br>(96.3%) | 60/61*<br>(98.4%) | 21/21*<br>(100%) |
| Patient Notification Rate | The proportion of participants completing the test procedure who received an immediate result notification | 44/46<br>(95.7%) | 262/267<br>(98.1%) | 79/82<br>(96.3%) | 120/122<br>(98.4%) | 63/63<br>(100%) |

\*The number of tests conducted is less than the number of samples collected, because samples were pooled into a single Xpert Ultra cartridge.

### Ethical considerations

This study was conducted according to the ethical principles set forth in the Declaration of Helsinki, ICH-GCP, European Directive 2001/20/EC, US Code of Federal Regulations Title 21, South African Good Clinical Practice Guidelines, and other local regulatory requirements. The study protocol was approved by the University of Pretoria Human Research Ethics Committee (HREC 391/2021) and by Boston University Institutional Review Board (H-44118).

## RESULTS

A total of 313 eligible HHCs where identified, of which 267/313 (85.3%) provided informed consent. All 267 participants completed the socio-demographics, clinical history and post-test acceptability survey. The median age of participants was 41 years (IQR 28-55). The majority were Black (87.6%, 234/267), 76.4% (204/267) were unemployed and 89.1% (238/267) had a monthly income of less than R5,000 ($268). A quarter (22.1%, 59/267) had a prior TB infection and just less than one fifth (16.9%, 45/267) were living with HIV. Almost a third of participants (30.3%, 81/267) screened positive for TB using the WHO-recommended four-symptom screener (W4SS).^39^

### Acceptability of In-Home, POC TUTT

Table 2 lists each of the eight acceptability constructs measured, the associated survey question, and its mean score. Of the 313 HHCs that met the study eligibility criteria, 267 (85.3%) provided informed consent, suggesting a high level of acceptability for in-home, POC TUTT, prior to being tested. The mean score for overall acceptability of in-home, POC TUTT across all eight acceptability constructs was 4.5 (SD=0.2), with 5 representing the highest level of acceptability. All constructs measuring a positive attitude towards in-home TB testing had mean scores above 4. The two constructs measuring negative attitudes toward in-home TB testing (ie., burden and opportunity cost), had mean scores of 1.4 and 1.3, respectively, with 1 representing the least negative response. When given the option to choose between sputum or tongue swabs for future in-home TB testing, 226/267 (84.6%) chose tongue swabs.

### Feasibility of Different Sputum and Tongue Swab-based In-home, POC Testing Methods

Table 3 lists and defines the four metrics used to assess feasibility and the associated performance of different testing methods against each. Tongue swab-based testing outperformed sputum testing on *Sample Collection Rate*. All 267 (100%) study participants were able to provide a tongue swab while only 17.2% (46/267) were able to successfully expectorate sputum for testing. Contact investigation teams successfully prepared all samples for immediate POC testing during a HCI, irrespective of the sample type or number of samples pooled. The *Test Processing Rate* was consequently 100% for both specimen types. The *Test Success Rate* for tongue swab-based tests (97.6%, 160/164) was comparable to sputum (95.7%, 44/46). Two sputum tests failed to provide a test result due to a testing-error (44/46) resulting in a *Test Success Rate* of 95.7%. Tongue swab-based tests had a 97.6% (160/164) *Test Success Rate* due to 3 tests returning an error result. All participants with actionable test results were immediately notified at the time of the household investigation. Due to a higher *Test Success Rate*, tongue swab-based tests had a higher *Patient Notification Rate* (98.1%) as 262 of the 267 participants received a valid in-home test result. Of the 46 participants able to provide a sputum, 44 (95.7%) received a valid, in-home test result. Among the various tongue swab-based testing methods, the pooling of three samples had a better outcome compared to both two-sample pooling and individual testing across all feasibility metrics.

## DISCUSSION

To our knowledge, The TB Home Study is the first to examine in-home collection and testing of sputum and tongue swabs for universal, POC TB testing during HCIs. Our initial proof-of-concept study reported that among HHCs with TB-related symptoms, in-home testing using sputum was both acceptable and feasible.^16,40^ The current findings demonstrate high acceptability for in-home POC TB testing among HHCs, irrespective of symptom presentation. In addition to sputum, the collection, pooling, and testing of tongue swab samples was highly feasible. Furthermore, the high collection yield of tongue swab specimens (100%) compared to sputum (17.2%) is a testament to the potential benefit of using less invasive sample types for TB testing if paired with highly sensitive backend molecular platforms. These results align with findings from previous research.^41–45^

The low yield of sputum specimens is consistent with previous studies highlighting a major shortcoming of sputum-based TB testing.^46^ Similar to concerns related to asymptomatic or pre-symptomatic cases being missed with symptom screening is the concern of patients who are missed who are symptomatic but unable to expectorate sputum, resulting in missed cases.^47^ There is growing evidence highlighting the potential of tongue swabs as a viable additional or alternative specimen for TB testing due to the increased sample yield and potentially higher probability of case detection.^48,49^ The acceptability of swab collection, highlighted during the COVID-19 pandemic, aligns with our findings, which showed high participant comfort and trust in the tongue swab-based testing process.^50^ A relatively low (30.3%, 81/267) proportion of HHCs presented with symptoms, highlighting the large proportion that would not have received further clinical evaluation under routine conditions. TUTT of high risk groups like HHCs could increase case detection by as much has 17%.^17^

While our findings support the acceptability and feasibility of in-home POC TUTT, other published work suggest that implementing rapid POC testing at scale could be challenging due to a myriad of logistical, operational, and resource constraints.^51,52^ Earlier research on rapid diagnostic tests for malaria has shown that contextual factors such as community trust, healthcare worker training, and resource availability can influence the successful implementation of POC testing.^24^ The accuracy of tongue swab testing for TB remains a concern.^53^ The optimal number of swabs and approach necessary to optimize DNA recovery during processing remains an active area of research.^54^ Similarly, although the pooling of sputum samples has been shown to be efficient at reducing costs and producing highly accurate results, uncertainty remains whether the same would hold true when pooling tongue swabs.^55^ While the sensitivity of tongue swabs for TB diagnosis remains variable, the specificity seems notably high, yielding and overall favorable diagnostic effect.^43^ These findings suggest that tongue swabs, in the absence of sputum, may serve as a suitable screening test for active TB disease, with a negative test result informing subsequent clinical decision making regarding eligibility for TB preventive treatment.

The importance of our findings is supported by a growing recognition for the need for accurate tests that enable prompt linkage to care, are implementable at POC, by healthcare workers with minimal training, and with results that are available in a single patient encounter.^56^ Consideration of the feasibility of collecting, processing, and testing specimens outside of a traditional clinical setting is essential to estimating the true potential of using alternative sample types for TB testing during community-based active case finding. Despite the introduction of several new platforms, gaps in the current TB diagnostic pipeline still remain.^57^ Exploring the potential of new platforms, assays, and testing methods that show promise for POC deployment across different use case scenarios plays an essential role in refining target product profiles.^53^ Findings from this research can be used to inform new use case development as well as to refine target product profiles aimed at delivering near-POC and POC TB testing.

A key strength of this study was the ability to use lay community healthcare workers to conduct HCIs and deliver in-home POC TUTT. South Africa continues to face a severe shortage of qualified health care workers, which has resulted in task shifting to a range of lay healthcare workers.^58^ The delivery of TB services by community healthcare workers has been shown to enhance population coverage, increase testing, improve early diagnosis and linkage to care.^59^ Another strength of the current analysis is the use of a theory-informed instrument to measure acceptability. No standardized or validated healthcare intervention acceptability instrument currently exists.^27^ Similar to most other evaluations, our previous work assessing the acceptability of in-home, POC TB testing relied on behavioral measures of acceptability such as study enrollment and/or dropout rate. However, several reasons other than low acceptability could explain why people decline or withdraw from a healthcare intervention, including lack of motivation, distrust, or privacy concerns. The TFA framework is innovative in that it provides conceptually distinct constructs that capture key dimensions of acceptability allowing the assessment of complex healthcare interventions.^37^

Our study had several limitations. First, the high level of acceptability of POC TB testing among HHCs might be an overestimation. The acceptability of TB testing might be higher among contacts of TB patients compared to the general population due to increased perceived risk and awareness of the disease. HHCs directly exposed to TB have a heightened understanding of the importance of early detection and treatment.^16^ Secondly, this analysis did not include a cost- or cost-effectiveness analysis to estimate and compare the difference in cost and outcomes of each testing method. Pooling tongue swabs into a single Xpert Ultra cartridge aims to reduce the total number of cartridges required, often cited as a significant factor driving testing costs.^60^ However, sputum remains the gold standard for TB testing due to its superior sensitivity over tongue swabs.^61^ Future studies should explore the cost-effectiveness of these different testing methods to weigh potential cost savings with decreasing test accuracy, assessing the “financial feasibility” and scalability of the proposed testing methods.^11^ Lastly, we only offered testing to HHCs older than 18 years of age. In 2022, over 1 million children under the age of 16 developed TB globally, with an estimated 225,000 children dying from TB and associated complications.^62^ In the same year, 7% of South Africa’s notified TB cases occurred in children age 14 years and younger.^63^ The majority of TB deaths in children occur from lack of diagnosis and treatment.^64^ Effective TB diagnosis in children is hampered by several factors, including the paucibacillary nature of TB, its shared symptoms with other common childhood diseases, and most significantly, difficulties collecting samples for testing.^65^ The collection of non-invasive samples like tongue swabs for TB screening among children during HCI combined with the delivery of TB preventative treatment could have a significant public health impact and be cost-effective in preventing TB deaths in South Africa.^66^ Failing to prioritize children and adolescents in future studies will continue to stifle progress towards TB targets.

## CONCLUSION

As novel platforms and diagnostics for decentralized molecular testing become more readily available, this study provides evidence to support their integration into existing strategies, including HCI. Furthermore, these findings provide support for the expansion of in-home TB testing by minimally trained lay healthcare workers. The ability to integrate molecular POC testing into community-based strategies can reduce the workload of already overburdened laboratory and clinical facilities, improve client satisfaction, and remove persistent barriers preventing equal access to services.^67^ In-home POC TUTT using either sputum or tongue swabs is highly acceptable and feasible. Rapid molecular TB testing with immediate result notification at POC reduces the burden placed on those at highest risk by offering testing services in a single consultation, improves access to testing, and shows great potential for early case detection and result notification.

## Data Availability

All data produced in the present study are available upon reasonable request to the authors

## Notes

### Competing Interest Statement

LL was supported by the National Institute of Mental Health of the National Institutes of Health under grant number K01MH119923. The content is solely the responsibility of the authors and does not necessarily represent the official views of the National Institutes of Health

### Clinical Trial

NCT04973371

### Funding Statement

Funding was provided by the United States National Institutes of Health (Grant # R01AI150485 and R21EB023679) and FIND.

## REFERENCES

1. WHO consolidated guidelines on tuberculosis. Module 3: diagnosis - rapid diagnostics for tuberculosis detection, 2021 update. Geneva: World Health Organization; 2021. Licence: CC BY-NC-SA 3.0 IGO

2. Thompson RR, Nalugwa T, Oyuku D, Tucker A, Nantale M, Nakaweesa A, et al. Multicomponent strategy with decentralised molecular testing for tuberculosis in Uganda: a cost and cost-effectiveness analysis. The Lancet Global Health. 2023 Feb 1;11(2):e278–86.

3. Global tuberculosis report 2023. Geneva: World Health Organization; 2023. Licence: CC BY-NC-SA IGO

4. Pai M, Dewan PK, Swaminathan S. Transforming tuberculosis diagnosis. Nat Microbiol. 2023 May;8(5):756–9.

5. Global tuberculosis report 2022. Geneva: World Health organization; 2022. licence: cc bY-Nc-sa 3.0 iGo.

6. Coleman S, Mahlangu H. The state of TB in South Africa. TB Accountability Consortium; 2024 Mar.

7. Shah HD, Nazli Khatib M, Syed ZQ, Gaidhane AM, Yasobant S, Narkhede K, et al. Gaps and Interventions across the Diagnostic Care Cascade of TB Patients at the Level of Patient, Community and Health System: A Qualitative Review of the Literature. Trop Med Infect Dis. 2022 Jul 15;7(7):136.

8. Medina-Marino A, Bezuidenhout D, Bezuidenhout C, Facente SN, Fourie B, Shin SS, et al. In-home TB Testing Using GeneXpert Edge is Acceptable, Feasible, and Improves the Proportion of Symptomatic Household Contacts Tested for TB: A Proof-of-Concept Study. Open Forum Infect Dis. 2024 May 13;11(6):ofae279.

9. de Vos L, Mazinyo E, Bezuidenhout D, Ngcelwane N, Mandell DS, Schriger SH, et al. Reasons for missed opportunities to screen and test for TB in healthcare facilities. Public Health Action. 2022 Dec 21;12(4):171–3.

10. Pf K C VS N A, M U, Mm C A MM. Estimating the magnitude of pulmonary tuberculosis patients missed by primary health care clinics in South Africa. The international journal of tuberculosis and lung disease□: the official journal of the International Union against Tuberculosis and Lung Disease [Internet]. 2018 Mar 1 [cited 2024 Oct 14];22(3). Available from: https://pubmed.ncbi.nlm.nih.gov/29471903/

11. Martinson NA, Nonyane BAS, Genade LP, Berhanu RH, Naidoo P, Brey Z, et al. Evaluating systematic targeted universal testing for tuberculosis in primary care clinics of South Africa: A cluster-randomized trial (The TUTT Trial). PLOS Medicine [Internet]. 2023 May [cited 2024 Oct 8];20(5). Available from: https://www.ncbi.nlm.nih.gov/pmc/articles/PMC10263318/

12. USAID Global TB Strategy, 2023–2030, Implementation Approach.

13. Sekandi J, Dobbin K, Oloya J, Okwera A, Whalen C, Corso P. Cost-Effectiveness Analysis of Community Active Case Finding and Household Contact Investigation for Tuberculosis Case Detection in Urban Africa. PLOS ONE. 2015 Feb 6;10:e0117009.

14. Bezuidenhout C, Long L, Nichols B, Meyer-Rath G, Fox MP, Theron G, et al. Sputum and tongue swab molecular testing for the in-home diagnosis of tuberculosis in unselected household contacts: a cost and cost-effectiveness analysis [Internet]. medRxiv; 2024 [cited 2024 Nov 1]. p. 2024.10.18.24315746. Available from: https://www.medrxiv.org/content/10.1101/2024.10.18.24315746v1

15. Andom AT, Gilbert HN, Ndayizigiye M, Mukherjee JS, Lively CT, Nthunya J, et al. Understanding barriers to tuberculosis diagnosis and treatment completion in a low-resource setting: A mixed-methods study in the Kingdom of Lesotho. PLoS One. 2023 May 11;18(5):e0285774.

16. Medina-Marino A, de Vos L, Bezuidenhout D, Denkinger CM, Schumacher SG, Shin SS, et al. “I got tested at home, the help came to me”: acceptability and feasibility of home-based TB testing of household contacts using portable molecular diagnostics in South Africa. Trop Med Int Health. 2021 Mar;26(3):343–54.

17. Berhanu RH, Lebina L, Nonyane BAS, Milovanovic M, Kinghorn A, Connell L, et al. Yield of Facility-based Targeted Universal Testing for Tuberculosis With Xpert and Mycobacterial Culture in High-Risk Groups Attending Primary Care Facilities in South Africa. Clinical Infectious Diseases. 2023 May 1;76(9):1594–603.

18. Kay A, Vasiliu A, Carratala-Castro L, Mtafya B, Reyes JEM, Maphalala N, et al. Performance of a stool-based quantitative PCR assay for the diagnosis of tuberculosis in adolescents and adults: a multinational, prospective diagnostic accuracy study. The Lancet Microbe. 2024 May 1;5(5):e433–41.

19. Insufficient quality of sputum submitted for tuberculosis diagnosis and associated factors, in Klaten district, Indonesia | BMC Pulmonary Medicine | Full Text [Internet]. [cited 2024 Aug 29]. Available from: https://bmcpulmmed.biomedcentral.com/articles/10.1186/1471-2466-9-16

20. Baik Y, Nakasolya O, Isooba D, Mukiibi J, Kitonsa PJ, Erisa KC, et al. Cost to perform door-to-door universal sputum screening for TB in a high-burden community. Int J Tuberc Lung Dis. 2023 Mar 1;27(3):195–201.

21. Steadman A, Andama A, Ball A, Mukwatamundu J, Khimani K, Mochizuki T, et al. New Manual Quantitative Polymerase Chain Reaction Assay Validated on Tongue Swabs Collected and Processed in Uganda Shows Sensitivity That Rivals Sputum-based Molecular Tuberculosis Diagnostics. Clin Infect Dis. 2024 May 15;78(5):1313–20.

22. Vuchas C, Teyim P, Dang BF, Neh A, Keugni L, Che M, et al. Implementation of large-scale pooled testing to increase rapid molecular diagnostic test coverage for tuberculosis: a retrospective evaluation. Sci Rep. 2023 Sep 16;13:15358.

23. Drain PK, Heichman KA, Wilson D. A new point-of-care test to diagnose tuberculosis. Lancet Infect Dis. 2019 Aug;19(8):794–6.

24. Beisel U, Umlauf R, Hutchinson E, Chandler CIR. The complexities of simple technologies: re-imagining the role of rapid diagnostic tests in malaria control efforts. Malaria Journal. 2016 Feb 5;15(1):64.

25. Chenai Mathabire Rücker S, Lissouba P, Akinyi M, Vicent Lubega A, Stewart R, Tamayo Antabak N, et al. Feasibility and acceptability of using the novel urine-based FujiLAM test to detect tuberculosis: A multi-country mixed-methods study. J Clin Tuberc Other Mycobact Dis. 2022 Apr 25;27:100316.

26. Medina-Marino A, Bezuidenhout D, Bezuidenhout C, Facente SN, Fourie B, Shin SS, et al. In-home TB Testing Using GeneXpert Edge is Acceptable, Feasible, and Improves the Proportion of Symptomatic Household Contacts Tested for TB: A Proof-of-Concept Study. Open Forum Infectious Diseases. 2024 Jun 3;11(6):ofae279.

27. Sekhon M, Cartwright M, Francis JJ. Development of a theory-informed questionnaire to assess the acceptability of healthcare interventions. BMC Health Services Research. 2022 Mar 1;22(1):279.

28. Pant Pai N, Chiavegatti T, Vijh R, Karatzas N, Daher J, Smallwood M, et al. Measures and Metrics for Feasibility of Proof-of-Concept Studies With Human Immunodeficiency Virus Rapid Point-of-Care Technologies. Point Care. 2017 Dec;16(4):141–50.

29. Buffalo City Metro EC. Profile and analysis district development model. Available at: https://www.cogta.gov.za/cgta_2016/wpcontent/uploads/2023/11/DistrictProfile_BUFFALOCITY07072020.pdf

30. South Africa, Department of Health. National-TB-Program-Strategic-Plan-2023-2028. Available at: https://tbthinktank.org/wp-content/uploads/2024/05/National-TB-Program-Strategic-Plan-2023-2028.pdf

31. National-TB-Program-Strategic-Plan-2023-2028.pdf [Internet]. [cited 2024 Oct 2]. Available from: https://tbthinktank.org/wp-content/uploads/2024/05/National-TB-Program-Strategic-Plan-2023-2028.pdf

32. District Health Barometer 2019/20. Health Systems Trust. Available at: https://www.hst.org.za/publications/District%20Health%20Barometers/DHB%20201920%20Complete%20Book.pdf. Accessed on 8 August 2024.

33. Andrew Medina-Marino, Dana Bezuidenhout, Charl Bezuidenhout, Shelley N Facente, Bernard Fourie, Sanghyuk S Shin, Adam Penn-Nicholson, Grant Theron, In-home TB Testing Using GeneXpert Edge is Acceptable, Feasible, and Improves the Proportion of Symptomatic Household Contacts Tested for TB: A Proof-of-Concept Study, Open Forum Infectious Diseases, Volume 11, Issue 6, June 2024, ofae279, 10.1093/ofid/ofae279

34. Bernard Fourie P. Community-based detection of M. tuberculosis from oral swab specimens [Internet]. Presentation presented at; St George’s, University of London; 2019 Oct 18 [cited 2024 Oct 14]. Available from: https://sgul.figshare.com/articles/presentation/Community-based_detection_of_M_tuberculosis_from_oral_swab_specimens/9970973/1

35. Daum LT, Fourie PB, Peters RPH, Rodriguez JD, Worthy SA, Khubbar M, et al. Xpert(®) MTB/RIF detection of Mycobacterium tuberculosis from sputum collected in molecular transport medium. Int J Tuberc Lung Dis. 2016 Aug;20(8):1118–24.

36. Mboneni TA, Eales OO, Maningi NE, Hugo JF, Fourie PB. Detection by RT-PCR of Mycobacterium tuberculosis from oral swab specimens using PrimeStore® molecular transport medium.

37. Sekhon M, Cartwright M, Francis JJ. Acceptability of healthcare interventions: an overview of reviews and development of a theoretical framework. BMC Health Services Research. 2017 Jan 26;17(1):88.

38. Pant Pai N, Chiavegatti T, Vijh R, Karatzas N, Daher J, Smallwood M, et al. Measures and Metrics for Feasibility of Proof-of-Concept Studies With Human Immunodeficiency Virus Rapid Point-of-Care Technologies: The Evidence and the Framework. Point of Care. 2017 Dec;16(4):141.

39. WHO consolidated guidelines on tuberculosis. Module 2: screening – systematic screening for tuberculosis disease. Geneva: World Health Organization; 2021. Licence: CC BY-NC-SA 3.0 IGO

40. Medina-Marino A, Bezuidenhout D, Bezuidenhout C, Facente SN, Fourie B, Shin SS, et al. In-home TB Testing Using GeneXpert Edge is Acceptable, Feasible and Improves the Proportion of Symptomatic Household Contacts Tested for TB: A Proof-of-Concept Study. Open Forum Infectious Diseases. 2024 May 13;ofae279.

41. Nooy A de, Ockhuisen T, Korobitsyn A, Khan SA, Ruhwald M, Ismail N, et al. Trade-offs between clinical performance and test accessibility in tuberculosis diagnosis: a multi-country modelling approach for target product profile development. The Lancet Global Health. 2024 Jul 1;12(7):e1139–48.

42. Wood RC, Luabeya AK, Dragovich RB, Olson AM, Lochner KA, Weigel KM, et al. Diagnostic accuracy of tongue swab testing on two automated tuberculosis diagnostic platforms, Cepheid Xpert MTB/RIF Ultra and Molbio Truenat MTB Ultima. Journal of Clinical Microbiology. 2024 Mar 14;62(4):e00019–24.

43. Church EC, Steingart KR, Cangelosi GA, Ruhwald M, Kohli M, Shapiro AE. Oral swabs with a rapid molecular diagnostic test for pulmonary tuberculosis in adults and children: a systematic review. Lancet Glob Health. 2023 Dec 12;12(1):e45–54.

44. Andama A, Whitman GR, Crowder R, Reza TF, Jaganath D, Mulondo J, et al. Accuracy of Tongue Swab Testing Using Xpert MTB-RIF Ultra for Tuberculosis Diagnosis. Journal of Clinical Microbiology. 2022 Jun 27;60(7):e00421–22.

45. Luabeya AK, Wood RC, Shenje J, Filander E, Ontong C, Mabwe S, et al. Noninvasive Detection of Tuberculosis by Oral Swab Analysis. J Clin Microbiol. 2019 Feb 27;57(3):e01847–18.

46. Ruhwald et al. Learning from COVID-19 to reimagine tuberculosis diagnosis. The Lancet Microbe, Volume 2, Issue 5, e169–e170

47. Stuck L, van Haaster AC, Kapata-Chanda P, Klinkenberg E, Kapata N, Cobelens F. How “Subclinical” is Subclinical Tuberculosis? An Analysis of National Prevalence Survey Data from Zambia. Clin Infect Dis. 2022 Sep 14;75(5):842–8.

48. Holtgrewe LML, Jain S, Dekova R, Broger T, Isaacs C, Theron G, Nahid P, Cattamanchi A, Denkinger CM, Yerlikaya S. Innovative COVID-19 Point-of-Care Diagnostics Suitable for Tuberculosis Diagnosis: A Scoping Review. Journal of Clinical Medicine. 2024; 13(19):5894. 10.3390/jcm13195894

49. Ahls C, David A, Shankar Chilambi G, Cattamanchi A, De Vos M, Heard K, et al. Xpert MTB/RIF Ultra testing from tongue swabs – Diluted SR method v1 [Internet]. 2024 [cited 2024 Sep 6]. Available from: https://www.protocols.io/view/xpert-mtb-rif-ultra-testing-from-tongue-swabs-dilu-dfjf3kjn

50. Yao H, Shen Y, Liang Z, Xue X, Zhao C, Xu X, et al. Superior effectiveness and acceptability of saliva samples for the detection of SARS-CoV-2 in China. Biosafety and Health. 2024 Apr 1;6(2):88–91.

51. Gavina K, Franco LC, Khan H, Lavik JP, Relich RF. Molecular point-of-care devices for the diagnosis of infectious diseases in resource-limited settings – A review of the current landscape, technical challenges, and clinical impact. Journal of Clinical Virology. 2023 Dec 1;169:105613.

52. Kuupiel D, Bawontuo V, Mashamba-Thompson TP. Improving the Accessibility and Efficiency of Point-of-Care Diagnostics Services in Low- and Middle-Income Countries: Lean and Agile Supply Chain Management. Diagnostics (Basel). 2017 Nov 29;7(4):58.

53. Nathavitharana RR, Garcia-Basteiro AL, Ruhwald M, Cobelens F, Theron G. Reimagining the status quo: How close are we to rapid sputum-free tuberculosis diagnostics for all? EBioMedicine. 2022 Apr;78:103939. doi: 10.1016/j.ebiom.2022.103939. Epub 2022 Mar 23. PMID: 35339423; PMCID: PMC9043971.

54. Nathavitharana RR, Garcia-Basteiro AL, Ruhwald M, Cobelens F, Theron G. Reimagining the status quo: How close are we to rapid sputum-free tuberculosis diagnostics for all. EBioMedicine. 2022 Apr 1;78:103939.

55. Cuevas LE, Santos VS, Lima SVMA, Kontogianni K, Bimba JS, Iem V, et al. Systematic Review of Pooling Sputum as an Efficient Method for Xpert MTB/RIF Tuberculosis Testing during the COVID-19 Pandemic. Emerg Infect Dis. 2021 Mar;27(3):719–27.

56. Kuupiel D, Bawontuo V, Mashamba-Thompson TP. Improving the Accessibility and Efficiency of Point-of-Care Diagnostics Services in Low- and Middle-Income Countries: Lean and Agile Supply Chain Management. Diagnostics (Basel). 2017 Nov 29;7(4):58.

57. Broger T, Marx FM, Theron G, Marais BJ, Nicol MP, Kerkhoff AD, et al. Diagnostic yield as an important metric for the evaluation of novel tuberculosis tests: rationale and guidance for future research. The Lancet Global Health. 2024 Jul 1;12(7):e1184–91.

58. Murphy JP, Moolla A, Kgowedi S, Mongwenyana C, Mngadi S, Ngcobo N, et al. Community health worker models in South Africa: a qualitative study on policy implementation of the 2018/19 revised framework. Health Policy and Planning. 2021 May 1;36(4):384–96.

59. Ajudua FI, Mash RJ. Implementing active surveillance for TB: A descriptive survey of healthcare workers in the Eastern Cape, South Africa. Afr J Prim Health Care Fam Med. 2024 Feb 23;16(1):4217.

60. MacLean ELH, Miotto P, González Angulo L, Chiacchiaretta M, Walker TM, Casenghi M, et al. Updating the WHO target product profile for next-generation Mycobacterium tuberculosis drug susceptibility testing at peripheral centres. PLOS Glob Public Health. 2023 Mar 31;3(3):e0001754.

61. Ealand CS, Sewcharran A, Peters JS, Gordhan BG, Kamariza M, Bertozzi CR, et al. The performance of tongue swabs for detection of pulmonary tuberculosis. Front Cell Infect Microbiol. 2023 Sep 6;13:1186191.

62. Global tuberculosis report 2021. Geneva: World Health Organization; 2021. Licence: CC BY-NC-SA 3.0 IGO.63. TB profile [Internet]. [cited 2024 Oct 14]. Available from: https://worldhealthorg.shinyapps.io/tb_profiles/?_inputs_&entity_type=%22country%22&iso2=%22ZA%22&lan=%22EN%22

64. Dodd PJ, Yuen CM, Sismanidis C, Seddon JA, Jenkins HE. The global burden of tuberculosis mortality in children: a mathematical modelling study. Lancet Glob Health. 2017 Sep;5(9):e898–906.

65. Wobudeya E, Bonnet M, Walters EG, Nabeta P, Song R, Murithi W, et al. Diagnostic Advances in Childhood Tuberculosis—Improving Specimen Collection and Yield of Microbiological Diagnosis for Intrathoracic Tuberculosis. Pathogens. 2022 Mar 23;11(4):389.

66. Brough J, Martinez L, Hatherill M, Zar HJ, Lo NC, Andrews JR. Public Health Impact and Cost-Effectiveness of Screening for Active Tuberculosis Disease or Infection Among Children in South Africa. Clinical Infectious Diseases. 2023 Dec 1;77(11):1544–51.

67. Saberi P, Ming K, Shrestha I, Scott H, Thorson B, Liu A. Feasibility and Acceptability of Home-Collected Samples for Human Immunodeficiency Virus Preexposure Prophylaxis and Severe Acute Respiratory Syndrome Coronavirus 2 Laboratory Tests in San Francisco Primary Care Clinics. Open Forum Infect Dis. 2022 Feb;9(2):ofab657.

